# South Africa’s male homicide epidemic hiding in plain sight: exploring sex differences and patterns in homicide risk in a retrospective descriptive study of postmortem investigations

**DOI:** 10.1101/2023.02.02.23285093

**Authors:** R Matzopoulos, M Prinsloo, S Mhlongo, L Marineau, M Cornell, B Bowman, TA Mamashela, N Gwebushe, A Ketelo, LJ Martin, B Dekel, C Lombard, R Jewkes, N Abrahams

## Abstract

**Background:** South Africa has homicide rates six times the global average, predominantly among men, but little is known about male victims. As part of the country’s first ever study of male homicide we compared 2017 male and female victim profiles for selected covariates, against global averages and previous estimates for 2009.

**Methods:** We conducted a retrospective descriptive study of routine data collected through postmortem investigations, calculating age-standardised mortality rates for manner of death by age, sex and province and male-to-female incidence rate ratios with 95% confidence intervals. We then used generalised linear models and linear regression models to assess the association between sex and victim characteristics including age and mechanism of injury (guns, stabs and blunt force) within and between years.

**Findings:** 87% of 19,477 homicides in 2017 were males, equating to seven male deaths for every female, with sharp force and firearm discharge the most common external causes. Rates were higher among males than females at all ages, and up to eight times higher among males aged 15-44 years. Provincial rates varied overall and by sex, with the highest comparative risk for men vs. women in the Western Cape Province (11.4 males for every 1 female). Male homicides peaked during December and were highest on weekends, underscoring the prominent role of alcohol as a risk factor. Significantly more males tested positive for alcohol than females.

**Interpretation:** The massive, disproportionate and enduring homicide risk borne by adult South African men highlights the negligible prevention response. Only through challenging the normative perception of male invulnerability can we begin to address the enormous burden of violence impacting men. There is an urgent need to address the insidious effect of such societal norms alongside implementing structural interventions to overcome the root causes of poverty and inequality and better control alcohol and firearms.

**Funding:** South African Medical Research Council and Ford Foundation.

## INTRODUCTION

In South Africa in 2009, injury-related mortality accounted for 8.6% of deaths(1) primarily due to extremely high homicide rates, which were nearly six times the global average.(2,3) Adult men, 20 years and older, accounted for more than three-quarters (79%) of all homicides in which the age of the decedent was known.(2) Despite this significant difference, there has been limited focus on the male victims of homicide. Two previous nationally representative homicide studies in 1999 and 2009 only explored the situational contexts of homicide for women and child victims.(4,5) This is consistent with global directives such as the 67^th^ World Health Assembly Resolution that have prioritised prevention efforts to reduce violence against women regardless of the far higher rates of violence against men. Globally men bear a far higher injury mortality and morbidity burden,(6) but we were unable to identify any studies that explored the different patterns of male and female homicide in South Africa.

To address this gap, the South African Medical Research Council (SAMRC) funded a comprehensive Female and Male Homicide and Injury Mortality Study (FAMHIS) for 2017. The first phase included a nationally representative all-cause injury mortality study. For homicide cases specifically, a second phase entailed the collection of more detailed information from interviews with police investigating officers. This replicates female homicide studies in 1999 and 2009,(4,5) but, for the first time, will provide comparable information describing the personal and situational risks for male homicide.

In this study, we analysed homicide data from the first phase of the study. We compared (1) male and female victim profiles by external cause, age, province, day of week, month and alcohol-relatedness and (2) male: female homicide rate ratios against global averages for selected covariates (external cause and age), and (3) explored whether the odds of male versus female homicide by external cause and age had changed between 2009 and 2017.

## METHODS

### Study design and data sources

We conducted a retrospective descriptive study of routine postmortem investigation data via a nationally representative survey of mortuaries sampled from eight of South Africa’s nine provinces for 2017. Data were obtained from postmortem reports and ancillary documentation, including police reports and hospital records. Survey data were combined with compatible routinely captured data from the Western Cape Province’s Forensic Pathology Service (FPS), which maintains full coverage of medico-legal mortuaries in the province.

### Sampling

We drew a multistage stratified cluster sample for eight provinces, using mortuaries as the primary sampling unit (cluster). We used a sampling frame of 58,641 postmortem reports from 121 mortuaries to draw a representative sample stratified by province and mortuary size: small (≤500 cases), medium (501–1500 cases) and large (>1500 cases). Forty-five mortuaries from eight provinces were selected with an expected sample of 22,733 records. To account for the selection probabilities of mortuaries within survey strata, we applied analysis weights. All records for the Western Cape were obtained from the provincial FPS. Further details on sampling, fieldwork and data collection methods are provided elsewhere(7).

### Case selection and variables

We excluded all deaths from natural causes, fetal deaths and deaths that occurred outside South Africa. For deaths due to external causes, we excluded suicide and deaths that were transport-related, or unintentional after redistributing deaths due to undetermined intent. For all homicides we ascribed an external cause of death consistent with the tenth revision of the International Statistical Classification of Diseases and Related Health Problems, 2007 (ICD-10; Supplementary Table 1). We defined weekends as Saturdays and Sundays and hot months as November through March. The mortuary death register number and death notification number were collected as identifiers for follow-up, and to resolve data capture errors, but were excluded from the analysis.

### Statistical analysis

We calculated age-standardised homicide rates (ASHR) by sex using 2017 population estimates provided by Dorrington (2013)(8) and the World Health Organization’s (WHO’s) world standard population, and male-to-female incidence rate ratios (IRRs) with 95% confidence intervals (CIs). Cases with unknown age were proportionally redistributed for males and females as follows:

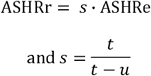

where ASHRr = ASHR with unknown age cases redistributed;

ASHRe = ASHR with unknown age cases excluded;

s = scaling factor; t = total number of homicides; u = homicides with unknown age

Similarly, we applied scaling factors to calculate adjusted homicide rates for comparison with the Global Burden of Disease study, which redistributes injury deaths due to undetermined intent to known causes, by proportionally distributing injury deaths ascribed to undetermined causes(7) to apparent manner of death (homicide, suicide and transport and other unintentional) by age and sex.

We used generalised linear models and linear regression models to assess the association between sex and victim characteristics including age and mechanism of injury (guns, stabs and blunt force) within and between years. Coefficients or relative risk (RR) and 95% CIs were reported. The models also included interaction terms between gender and year to compare males’ and females’ homicide characteristics between the two years; p values were reported and associations assessed using a significance level of alpha = 0.05.

### Ethics

Ethical approval for the study was granted by the Ethics Committee of the SAMRC (EC 008-5-2018). Further approval and access to data were obtained from the National and Provincial Departments of Health and Forensic Pathology Service.

## RESULTS

Our total sample of 22,822 injury cases exceeded the expected sample by 89 cases. After applying sampling weights we estimated a total of 54,734 injury deaths (data not shown), and 19,477 homicides of which 87% were male (Table 1). Men had a much higher age standardised homicide rate than women (59.7 vs. 9.0 per 100,000 population), equivalent to 7 male deaths for every 1 female death.

**Table 1.** Descriptive male and female homicide victim characteristics in South Africa in 2017 by external cause of death, age, province, population group, month of year, day of week and alcohol-relatedness (weighted)

|  | Male |  |  | M/F Incidence<br>Rate Ratio<br>(95% CI) | Female |  |  |
| --- | --- | --- | --- | --- | --- | --- | --- |
|  | Number<br>(95%CI) | Percentage<br>(95%CI)** | Age-standardised<br>rate/100 000<br>population (95%CI)* |  | Number<br>(95%CI) | Percentage (95%CI)** | Age-standardised<br>rate/100 000<br>population (95%CI)* |
| **All homicides (n=19477)\*** | **16835 (15735, 17936)** | **86.7 (86.2, 87.2)** | **59.7 (55.5, 63.9)** | **6.9 (6.4, 7.4)** | **2583 (2351, 2814)** | **13.3 (12.8, 13.8)** | **9.0 (7.8, 10.1)** |
| <u>External cause (n=19477)</u> | 16835 (15735, 17936) |  |  |  | 2583 (2351, 2814) |  |  |
| Sharp force | 7071 (6560, 7582) | 42.0 (40.5, 43.5) | 24.4 (22.3, 26.6) | 8.4 (7.5, 9.5) | 885 (771, 999) | 34.3 (32.5, 36.1) | 3.1 (2.4, 3.7) |
| Firearm discharge | 5616 (4955, 6278) | 33.4 (31.3, 35.5) | 20.1 (17.4, 22.9) | 9.0 (7.8, 10.3) | 659 (609, 708) | 25.5 (23.9, 27.2) | 2.3 (1.9, 2.7) |
| Blunt force | 3111 (2940, 3282) | 18.5 (17.0, 20.1) | 11.3 (10.2, 12.4) | 6.0 (5.1, 7.0) | 546 (496, 595) | 21.1 (19.8, 22.6) | 1.9 (1.5, 2.3) |
| Strangled/asphyxiated/suffocated | 212 (183, 241) | 1.3 (1.1, 1.4) | 0.8 (0.6, 1.1) | 1.0 (0.7, 1.3) | 232 (201, 263) | 9.0 (8.1, 10.0) | 0.8 (0.5, 1.1) |
| Fire /other burn | 125 (90, 160) | 0.7 (0.6, 1.0) | 0.5 (0.2, 0.8) | 3.1 (1.7, 5.5) | 43 (32, 55) | 1.7 (1.2, 2.3) | 0.2 (0.1, 0.2) |
| Other*** | 180 (98, 262) | 1.1 (0.7, 1.6) | 0.6 (0.2, 1.0) | 1.6 (1.1, 2.3) | 122 (77, 167) | 4.7 (3.5, 6.4) | 0.4 (0.3, 0.5) |
| Unknown**** | 520 (435, 606) | 3.1 (2.5, 3.8) | 3.0 (0.8, 5.3) | 5.7 (3.9, 8.2) | 96 (74, 118) | 3.7 (2.9, 4.7) | 0.3 (0.1, 0.6) |
| <u>Age in years (n=18984)</u> | 16465 (15513, 17416) |  |  |  | 2506 (2304, 2708) |  |  |
| 0-4 | 153 (102, 204) | 0.9 (0.7, 1.2) | 5.3 (4.4, 6.1) | 1.8 (1.1, 2.8) | 84 (47, 121) | 3.4 (2.3, 4.9) | 3.0 (2.3, 3.6) |
| 5-14 | 122 (101, 143) | 0.7 (0.6, 0.9) | 2.3 (1.9, 2.8) | 1.7 (1.1, 2.8) | 70 (56, 85) | 2.8 (2.3, 3.4) | 1.4 (1.0, 1.7) |
| 15-29 | 7621 (7184, 8058) | 46.3 (45.6, 47.0) | 101.2 (98.9, 103.4) | 8.4 (7.4, 9.4) | 898 (817, 979) | 35.8 (34.4, 37.3) | 12.1 (11.3, 12.9) |
| 30-44 | 6012 (5595, 6429) | 36.5 (35.9, 37.2) | 93.0 (90.6, 95.4) | 7.9 (6.9, 9.0) | 760 (673, 846) | 30.3 (28.9, 32.0) | 11.8 (10.9, 12.6) |
| 45-59 | 1894 (1775, 2012) | 11.5 (10.9, 12.1) | 55.4 (52.9, 57.9) | 5.9 (4.9, 7.0) | 387 (338, 437) | 15.5 (13.6, 17.5) | 9.5 (8.5, 10.4) |
| 60-69 | 461 (417, 505) | 2.8 (2.5, 3.1) | 37.3 (33.9, 40.7) | 3.8 (2.8, 5.2) | 162 (142, 182) | 6.5 (5.6, 7.4) | 9.8 (8.3, 11.3) |
| 70-79 | 163 (137, 190) | 1.0 (0.8, 1.2) | 30.2 (25.6, 34.8) | 3.3 (2.1, 5.2) | 79 (64, 93) | 3.1 (2.7, 3.7) | 9.1 (7.1, 11.1) |
| 80+ | 39 (25, 52) | 0.2 (0.2, 0.3) | 19.1 (13.1, 25.1) | 1.3 (0.7, 2.6) | 66 (47, 84) | 2.6 (1.9, 3.6) | 14.5 (11.0, 18.1) |
| Mean age (SD) | 32.5 (12.7) |  |  |  | 36.4 (17.5) |  |  |
| <u>Province (n=19477)</u> | 16835 (15735, 17936) |  |  |  | 2583 (2351, 2814) |  |  |
| Eastern Cape | 2841 (2667, 3016) | 16.9 (15.5, 18.3) | 97.2 (85.6, 108.8) | 5.4 (4.7, 6.3) | 599 (541, 657) | 23.2 (20.6, 26.0) | 17.5 (12.0, 22.9) |
| Free State | 960 (819, 1100) | 5.7 (4.9, 6.6) | 69.5 (54.9, 84.2) | 7.2 (5.3, 9.7) | 143 (111, 176) | 5.6 (4.4, 7.0) | 10.0 (5.8, 14.2) |
| Gauteng | 3812 (2859, 4764) | 22.6 (18.5, 27.4) | 47.0 (29.9, 64.1) | 6.9 (5.9, 8.0) | 539 (346, 731) | 20.9 (15.5, 27.5) | 7.0 (2.7, 11.4) |
| Kwazulu Natal | 3703 (3268, 4138) | 22.0 (19.7, 24.5) | 73.6 (60.9, 86.3) | 6.7 (5.8, 7.7) | 608 (512, 705) | 23.6 (20.3, 27.2) | 10.9 (8.2, 13.7) |
| Limpopo | 640 (465, 815) | 3.8 (2.9, 5.0) | 27.0 (14.8, 39.3) | 6.8 (4.8, 9.7) | 108 (70, 146) | 4.2 (3.1, 5.6) | 3.7 (0.8, 6.6) |
| Mpumalanga | 581 (469, 693) | 3.5 (2.8, 4.2) | 26.7 (18.7, 34.7) | 5.2 (3.7, 7.2) | 118 (91, 145) | 4.6 (3.7, 5.7) | 5.1 (2.5, 7.8) |
| Northern Cape | 211 (107, 314) | 1.3 (0.8, 2.0) | 39.2 (0.0, 80.6) | 4.7 (2.7, 8.1) | 46 (23, 70) | 1.8 (1.1, 3.0) | 8.7 (0.0, 20.0) |
| Northwest | 628 (517, 739) | 3.7 (3.1, 4.5) | 32.2 (19.7, 44.7) | 5.8 (4.1, 8.2) | 105 (102, 108) | 4.1 (3.7, 4.5) | 5.8 (3.4, 8.2) |
| Western Cape | 3460 (3460, 3460) | 20.6 (19.2, 21.9) | 100.7 (100.7, 100.7) | 11.4 (9.4, 13.9) | 316 (316, 316) | 12.2 (11.1, 13.4) | 9.2 (8.8, 9.5) |
| <u>Month of year (n=19443)</u> | 16808 (15708, 17909) |  |  |  | 2582 (2350, 2813) |  |  |
| January | 1170 (1104, 1236) | 7.0 (6.7, 7.3) | 4.3 (3.7, 4.8) | 6.7 (5.2, 8.8) | 183 (143, 224) | 7.1 (5.4, 9.3) | 0.6 (0.5, 0.8) |
| February | 1168 (1088, 1248) | 6.9 (6.7, 7.2) | 4.4 (3.9, 4.9) | 6.7 (5.1, 8.7) | 184 (157, 211) | 7.1 (6.2, 8.1) | 0.6 (0.4, 0.9) |
| March | 1378 (1286, 1471) | 8.2 (7.7, 8.7) | 5.1 (4.3, 5.9) | 6.7 (5.3, 8.6) | 215 (193, 237) | 8.3 (7.5, 9.2) | 0.8 (0.5, 1.0) |
| April | 1579 (1480, 1679) | 9.4 (9.0, 9.8) | 5.7 (5.0, 6.4) | 6.3 (5.1, 7.9) | 263 (220, 307) | 10.2 (9.0, 11.5) | 0.9 (0.6, 1.2) |
| May | 1266 (1152, 1380) | 7.5 (7.2, 7.9) | 4.7 (3.9, 5.6) | 5.7 (4.5, 7.2) | 234 (177, 291) | 9.1 (7.6, 10.8) | 0.8 (0.5, 1.1) |
| June | 1205 (1072, 1339) | 7.2 (6.8, 7.6) | 4.4 (3.7, 5.1) | 7.0 (5.4, 9.1) | 181 (135, 227) | 7.0 (5.8, 8.4) | 0.6 (0.4, 0.8) |
| July | 1514 (1359, 1670) | 9.0 (8.6, 9.5) | 5.7 (4.8, 6.7) | 7.0 (5.5, 8.9) | 227 (192, 263) | 8.8 (8.0, 9.6) | 0.8 (0.5, 1.0) |
| August | 1264 (1178, 1351) | 7.5 (7.3, 7.8) | 4.6 (4.0, 5.3) | 7.2 (5.5, 9.3) | 185 (149, 222) | 7.2 (6.2, 8.3) | 0.6 (0.5, 0.8) |
| September | 1519 (1403, 1635) | 9.0 (8.7, 9.4) | 5.6 (4.7, 6.5) | 7.2 (5.7, 9.2) | 221 (201, 241) | 8.6 (7.9, 9.3) | 0.8 (0.5, 1.0) |
| October | 1395 (1310, 1480) | 8.3 (7.9, 8.7) | 5.3 (4.4, 6.2) | 7.2 (5.6, 9.2) | 205 (174, 235) | 7.9 (7.2, 8.7) | 0.7 (0.4, 1.0) |
| November | 1455 (1326, 1585) | 8.7 (8.3, 9.0) | 5.4 (4.6, 6.2) | 7.0 (5.5, 8.9) | 219 (197, 241) | 8.5 (7.7, 9.3) | 0.8 (0.5, 1.0) |
| December | 1894 (1756, 2032) | 11.3 (10.9, 11.7) | 7.0 (6.0, 7.9) | 7.6 (6.1, 9.4) | 264 (239, 289) | 10.2 (9.1, 11.5) | 0.9 (0.7, 1.2) |
| <u>Day of week (n=19443)</u> | 16808 (15708, 17909) |  |  |  | 2582 (2350, 2813) |  |  |
| Monday | 2123 (1979, 2267) | 12.6 (12.2, 13.1) | 8.0 (6.9, 9.1) | 5.6 (4.7, 6.8) | 397 (341, 453) | 15.4 (14.2, 16.6) | 1.4 (1.0, 1.7) |
| Tuesday | 1640 (1477, 1803) | 9.8 (9.3, 10.3) | 6.3 (5.2, 7.3) | 5.2 (4.3, 6.4) | 331 (284, 379) | 12.8 (11.8, 13.9) | 1.2 (0.8, 1.5) |
| Wednesday | 1564 (1452, 1676) | 9.3 (8.7, 9.9) | 5.9 (5.0, 6.8) | 5.1 (4.2, 6.3) | 320 (285, 356) | 12.4 (10.9, 14.1) | 1.1 (0.7, 1.5) |
| Thursday | 1505 (1378, 1631) | 9.0 (8.7, 9.2) | 5.9 (5.0, 6.8) | 5.7 (4.6, 7.1) | 279 (235, 324) | 10.8 (9.8, 12.0) | 1.0 (0.6, 1.3) |
| Friday | 1918 (1751, 2084) | 11.4 (11.0, 11.8) | 7.2 (6.1, 8.2) | 8.2 (6.5, 10.2) | 247 (217, 277) | 9.6 (8.7, 10.5) | 0.9 (0.6, 1.2) |
| Saturday | 3763 (3508, 4018) | 22.4 (21.7, 23.1) | 13.6 (11.9, 15.2) | 9.3 (7.9, 11.1) | 424 (356, 491) | 16.4 (14.9, 18.1) | 1.5 (1.1, 1.8) |
| Sunday | 4296 (4005, 4588) | 25.6 (24.9, 26.2) | 15.5 (14.0, 16.9) | 7.8 (6.7, 9.0) | 583 (531, 635) | 22.6 (20.5, 24.8) | 2.0 (1.5, 2.5) |
| <u>Blood Alcohol Concentration (n=3363)</u> |  |  |  |  |  |  |  |
| Positive BAC | 1626 (1152, 2099) | 54.8 (53.2, 56.3) | 5.6 (4.2, 7.0) | 11.4 (8.6, 15.2) | 150 (89, 211) | 38.2 (37.1, 39.3) | 0.5 (0.3, 0.7) |
| Mean positive g/100ml (SD; range) | 0.09 (0.10; 0.00, 0.49) |  |  |  | 0.06 (0.08, 0.00, 0.46) |  |  |

\* Column totals do not always sum to *n* due to 59 cases where sex was recorded as 'undetermined'. All rates, except rates for age in years, are age standardised.
\*\* Row percentages are displayed for "All homicide" and column percentages for covariates.
\*\*\* Includes, for males: neglect and abandonment (102 cases), poisoning (45 cases), being pushed from a height (11 cases), drowning/ immersion (9 cases), crushing (2 cases), electrocution (3 cases), and assault by other specified means 8 cases). For females: neglect and abandonment (70 cases), poisoning (21 cases), drowning/ immersion (4 cases), being pushed from a height (2 cases), electrocution (2 cases), maternal deaths/abortion related (5 cases), and assault by other specified means (18 cases).
\*\*\*\* Unknown includes assault by other unspecified means.

The most common external causes of death were sharp force, firearm discharge and blunt force injuries, with significantly higher rates among men than women for these causes (24.4, 20.1 and 11.3 vs 3.1, 2.3 and 1.9/100,000 respectively). A higher proportion of women than men died due to strangulation or asphyxiation (8.9% vs 1.2%), but rates were equivalent.

Men had far higher homicide rates than women in all age groups, with the highest rates in the 15-29 (101.2 vs 12.1) and 30-44 year age groups (93.0 vs 11.8)/100,00 population, equating to 8.4 and 7.9 male deaths for every female death in these age groups respectively.

There was considerable interprovincial variation by sex: three to four times higher in the provinces with the highest homides rates compared to provinces with the lowest rates among both males and females. For males age-standardised homicide rates ranged from 26.7 in Mpumalanga to 100.7/100,000 in the Western Cape; for females the lowest homicide rates were recorded in Limpopo (3.7 per 100,000 population) and the highest in the Eastern Cape (17.5). The male age-standardised homicide rates in the Western Cape were significantly higher than all provinces except the Eastern Cape, which were in turn significantly higher than all other provinces except KwaZulu-Natal. The highest male:female incidence rate ratio (IRR) was recorded in the Western Cape with 11.4 males for every female homicide.

There was also considerable interprovincial variation in the external cause of homicide by sex, particularly for firearm homicides, 88% of which occurred in four provinces (Eastern Cape, KwaZulu Natal, Gauteng and the Western Cape). Sharp force injuries were the leading cause of male homicide in all provinces except Gauteng and the Western Cape, where firearms were the leading cause. For females, sharp force was the leading cause of homicide in all provinces except Mpumalanga.

Temporally, both sexes followed a similar pattern with the highest percentage of cases coinciding with festive periods – December (Christmas) and April (Easter) – and school holidays – July and September. Close to half of all homicides were recorded on weekend days compared with weekdays. Disproportionately more men than women were murdered on week and weekend days, particularly on Saturdays (9.3 male deaths for every 1 female death). Rates on Mondays were higher than on other week days. A significantly higher percentage of male homicide victims tested positive for blood alcohol than females (11.4 males for 1 female).

Overall homicide rates could be more than 12% higher than shown in Table 1. Adjusting the age standardised and age specific rates by apportioning additional injury deaths of undetermined intent (Table 2), the overall adjusted homicide rate in South Africa was 7.1 times the global average, and higher among males than females (7.4 vs 5.9 times the global average respectively). There was considerable variation by external cause and age. Notably, IRRs for sharp force injuries amongst males and females were considerably higher than global averages (12.0 and 7.4 times respectively). For males, the highest IRRs were recorded amongst young adults aged 25-34 years, with homicide rates over eight times the global average. For females IRRs were highest in the older age categories, peaking at 10.9 among women older than 70 years.

**Table 2.** Male and female homicide rates in South Africa, 2017 (weighted), compared to global rates from the Global Burden of Disease Study, 2017,[12] by external cause of death and age.

|  | Male |  |  |  | Female |  |  |  | Overall |  |  |  |
| --- | --- | --- | --- | --- | --- | --- | --- | --- | --- | --- | --- | --- |
|  | IMS | IMS Adjusted * | GBD Global | IMS Adjusted:<br>GBD Global<br>Incidence Rate<br>Ratio | IMS | IMS Adjusted | GBD Global | IMS Adjusted:<br>GBD Global<br>Incidence Rate<br>Ratio | IMS | IMS Adjusted | GBD Global | IMS Adjusted:<br>GBD Global<br>Incidence Rate<br>Ratio |
|  | Age-standardised rate/100 000 population (95%CI) |  |  | Ratio | Age-standardised rate/100 000 population (95%CI) |  |  | Ratio | Age-standardised rate/100 000 population (95%CI) |  |  | Ratio |
| All homicide (n=19477) | **59.7 (55.5, 63.9)** | **66.0 (60.8, 71.2)** | **8.9 (8.4, 9.5)** | **7.4** | **9.0 (7.8, 10.1)** | **10.6 (9.1, 11.4)** | **1.8 (1.7, 2.0)** | **5.9** | **34.0 (31.6, 36.4)** | **38.2 (35.2, 41.2)** | **5.4 (5.1, 5.7)** | **7.1** |
| Firearm discharge | 20.1 (17.4, 22.9) | 22.2 (19.1, 25.5) | 4.2 (4.0, 4.4) | 5.3 | 2.3 (1.9, 2.7) | 2.7 (2.2, 3.1) | 0.5 (0.5, 0.5) | 5.4 | 11.0 (9.6, 12.4) | 12.4 (10.7, 14.0) | 2.3 (2.2, 2.5) | 5.4 |
| Sharp force | 24.4 (22.3, 26.6) | 27.0 (24.4, 29.6) | 2.1 (1.7, 2.3) | 12.9 | 3.1 (2.4, 3.7) | 3.7 (2.8, 4.2) | 0.5 (0.4, 0.5) | 7.4 | 13.9 (12.6, 15.2) | 15.6 (14.0, 17.2) | 1.3 (1.1, 1.4) | 12.0 |
| Other** | 15.1 (13.4, 16.8) | 16.7 (14.7, 18.7) | 2.6 (2.4, 2.9) | 6.4 | 3.6 (3.0, 4.3) | 4.2 (3.5, 4.9) | 0.9 (0.8, 1.0) | 4.7 | 9.3 (8.4, 10.2) | 10.5 (9.4, 11.5) | 1.8 (1.6, 1.9) | 5.8 |
|  | Age-specific rate/ 100 000 population (95% CI) |  |  | Ratio | Age-specific rate/ 100 000 population (95% CI) |  |  | Ratio | Age-specific rate/ 100 000 population (95% CI) |  |  | Ratio |
| Age in years (n=18984) |  |  |  |  |  |  |  |  |  |  |  |  |
| 0-14 | 3.4 (3.0, 3.8) | 4.1 (3.6, 4.6) | 1.5 (1.2, 1.8) | 2.7 | 1.9 (1.6, 2.2) | 2.5 (2.1, 2.9) | 1.1 (0.9, 1.3) | 2.3 | 2.7 (2.4, 2.9) | 3.4 (3.0, 3.6) | 1.3 (1.1, 1.5) | 2.6 |
| 15-19 | 49.8 (46.9, 52.7) | 52.1 (48.7, 55.5) | 10.9 (9.9, 12.0) | 4.8 | 7.0 (5.9, 8.0) | 8.2 (6.8, 9.5) | 1.8 (1.7, 2.1) | 4.6 | 28.2 (26.7, 29.8) | 30.3 (28.5, 32.3) | 6.5 (5.9, 7.1) | 4.7 |
| 20-24 | 120.4 (116.1, 124.7) | 126.7 (121.5, 131.8) | 17.8 (16.7, 19.0) | 7.1 | 12.9 (11.5, 14.3) | 14.7 (12.9, 16.5) | 2.4 (2.2, 2.7) | 6.1 | 66.3 (64.0, 68.5) | 70.7 (67.9, 73.4) | 10.2 (9.6, 10.9) | 6.9 |
| 25-29 | 131.5 (127.1, 135.8) | 139.6 (134.3, 144.8) | 16.3 (15.3, 17.4) | 8.6 | 15.3 (13.8, 16.7) | 17.3 (15.3, 19.1) | 2.3 (2.1, 2.6) | 7.5 | 72.9 (70.7, 75.2) | 78.1 (75.4, 80.9) | 9.4 (8.8, 10.0) | 8.3 |
| 30-34 | 114.1 (109.9, 118.2) | 121.9 (116.8, 126.8) | 15.3 (14.4, 16.3) | 8.0 | 13.0 (11.6, 14.3) | 14.8 (13.0, 16.5) | 2.4 (2.2, 2.6) | 6.2 | 63.1 (60.9, 65.2) | 68.1 (65.4, 70.7) | 8.9 (8.4, 9.5) | 7.7 |
| 35-39 | 88.1 (84.0, 92.1) | 96.7 (91.6, 101.8) | 14.1 (13.3, 15.0) | 6.9 | 11.0 (9.6, 12.5) | 12.6 (10.8, 14.6) | 2.3 (2.1, 2.5) | 5.5 | 49.2 (47.1, 51.4) | 54.4 (51.8, 57.2) | 8.2 (7.8, 8.8) | 6.6 |
| 40-44 | 69.1 (65.3, 73.0) | 74.8 (70.2, 79.6) | 12.0 (11.4, 12.9) | 6.2 | 10.8 (9.2, 12.3) | 12.3 (10.3, 14.3) | 2.2 (2.1, 2.4) | 5.6 | 40.6 (38.4, 42.7) | 44.4 (41.7, 47.0) | 7.2 (6.8, 7.6) | 6.2 |
| 45-49 | 56.9 (53.1, 60.6) | 63.3 (58.5, 68.0) | 10.0 (9.4, 10.6) | 6.3 | 10.9 (9.1, 12.6) | 12.2 (10.0, 14.4) | 2.0 (1.9, 2.1) | 6.1 | 35.4 (33.2, 37.5) | 39.5 (36.7, 42.2) | 6.0 (5.7, 6.4) | 6.6 |
| 50-69 | 35.3 (33.5, 37.1) | 40.6 (38.3, 43.0) | 7.8 (7.4, 8.2) | 5.2 | 12.2 (11.0, 13.4) | 14.4 (12.8, 16.0) | 1.9 (1.8, 2.0) | 7.6 | 25.1 (24.0, 26.3) | 29.1 (27.7, 30.7) | 4.8 (4.6, 5.0) | 6.1 |
| 70+ | 15.4 (13.3, 17.5) | 18.2 (15.4, 21.0) | 5.0 (4.7, 5.4) | 3.6 | 19.4 (16.2, 22.5) | 22.9 (18.8, 27.0) | 2.1 (1.9, 2.2) | 10.9 | 16.8 (15.0, 18.6) | 19.8 (17.5, 22.2) | 3.4 (3.2, 3.6) | 5.8 |
| Mean age (SD) | 32.5 (12.7) |  |  |  | 36 (17.5) |  |  |  | 33.0 (13.6) |  |  |  |

\* Adjusted for undetermined cause of unnatural death as presented in Prinsloo et al (2021).
\*\* Includes blunt force, strangled/asphyxiated/suffocated, being pushed from a height, drowning/immersion, poisoning from ingestion, poisoning from gas, fire or other burn, neglect and abandonment, maternal death/ abortion related, crushing, electrocution, assault by other specified means, and unknown cases.

Male victims were on average four years younger than females in 2009 and 2017 (Table 3).

**Table 3.** Comparison of homicide characteristics between 2009 and 2017 by sex and effect measure of study year and sex.

| Characteristic | Male |  | Female |  | Effect measure of study year and sex |  | p value |
| --- | --- | --- | --- | --- | --- | --- | --- |
|  | 2009 | 2017 | 2009 | 2017 | Sex, RR (95% CI) |  |  |
| Age, median (IQR) | 29.0 (23.0, 39.0) | 30.0 (24.0, 38.0) | 34.0 (23.0, 47.0) | 32.0 (24.0, 47.0) | Female: 1.00<br>Male: -4.08 (-4.92, -3.24)© | Female: 1.00<br>Male: -3.84 (-4.84, -2.83)© | 0.711 |
| Died from sharp force injuries, percent (95% CI) | 43.8 (41.1, 46.5) | 42.0 (40.5, 43.5) | 30.0 (27.7, 32.5) | 34.2 (32.5, 36.1) | Female: 1.00<br>Male: 1.46 (1.36, 1.56) | Female: 1.00<br>Male: 1.23 (1.16, 1.30) | <0.001 |
| Died from gunshot injuries, percent (95% CI) | 30.1 (27.6, 32.7) | 33.4 (31.3, 35.5) | 22.3 (20.0, 24.9) | 25.5 (23.9, 27.2) | Female: 1.00<br>Male: 1.35 (1.27, 1.44) | Female: 1.00<br>Male: 1.31 (1.17, 1.46) | 0.613 |
| Died from blunt force injuries, percent (95% CI) | 22.1 (20.9, 23.5) | 18.5 (17.0, 20.1) | 26.8 (24.8, 29.0) | 21.1 (19.7, 22.5) | Male: 1.00<br>Female: 1.21 (1.12, 1.31) | Male: 1.00<br>Female: 1.14 (1.04, 1.24) | 0.295 |
| Died on a weekend, percent (95% CI) | 46.0 (44.9, 47.1) | 48.0 (46.8, 49.1) | 37.6 (35.5, 39.7) | 39.0 (37.1, 40.9) | Female: 1.00<br>Male: 1.22 (1.16, 1.29) | Female: 1.00<br>Male: 1.23 (1.17, 1.29) | 0.885 |
| Died during hot season, percent (95% CI) | 42.9 (41.8, 44.0) | 42.0 (41.4, 42.7) | 42.7 (40.6, 44.8) | 41.3 (37.3, 45.4) | Female: 1.00<br>Male: 1.01 (0.96, 1.05) | Female: 1.00<br>Male: 1.02 (0.93, 1.12) | 0.788 |

© coefficient

Males had a significantly higher risk of being killed by sharp force than females in both 2009 and 2017 [RR=1.46 (9% CI: 1.36, 1.56) vs RR=1.23 (95% CI: 1.16, 1.46)] respectively. This represented a significant decrease in risk for males and a corresponding increase for females between years. The risk of dying from gunshot injuries increased for both men and women in this period, and was higher for men than women in both 2009 and 2017 [RR=1.35 (9% CI: 1.27, 1.44) vs RR=1.31 (95% CI: 1.17, 1.46) respectively. Female risk was higher than men for blunt force injuries [RR=1.21 (9% CI: 1.12, 1.31) in 2009 vs RR=1.14 (95% CI: 1.04, 1.24) in 2017], with no change between years. Males had a significantly higher risk of dying on weekends than females in both years [RR=1.22 (9% CI: 1.16, 1.29) vs RR=1.23 (95% CI: 1.17, 1.29)] respectively, with no change over time. There were no significant differences in homicide risk during the hot season by sex or year.

## DISCUSSION

These findings confirm the enduring nature of South Africa’s interpersonal violence problem in 2017 and the massive, disproportionate homicide risk borne by adult men. The disproportionate burden of homicide borne by South African men has already been shown in previous national estimates for 2000 in which males accounted for 84% of homicides,(10) and in 2009 (86%).(2) The situation continues to deteriorate in South Africa, consistent with global data showing that men are accounting for an increasing share of homicide,(11) but at a far greater magnitude locally. The disaggregated homicide pattern presented in this study is similar to countries in Latin America and the Caribbean with high overall homicide rates (>25 per 100,000 population) and an excessively high male proportion (>80%). Conversely, countries with low homicide rates <5 per 100,000 population have a lower proportion (<60%) of male homicides.(6) The fact that men are both perpetrators and victims of homicides masks the understanding that men are extremely vulnerable in many contexts. Yet responding to this inequity impacting men is complicated by the greater power men frequently hold in high violence settings and public health responses to addressing violence continue to focus on women and children.

Although the risk of homicide was higher for men than women at all ages, age strongly predicted the risk of homicide, with victims predominantly males 15-44 years old, and the sex disparity starting at a young age. A previous national survey reported a five times higher homicide rate among boys than girls.(12) The risk factors for interpersonal violence in South Africa are well understood.(17) However, the plethora of co-occurring risks exacerbates the risk of violence in South Africa, as it does in other high violence settings where the risk of violence is consistently greater in areas of lower socioeconomic status and where there are greater economic disparities and where there are legacies of colonialism, migrant labour, slavery, other forms of discrimination and human rights violations.

Whereas demographics also determine risk for homicide globally – highest among young adults and males – these risks in South Africa are compounded by socioeconomic factors with high concentrations of homicide in extremely poor neighbourhoods.(15) The country is among the world’s most unequal,(16) a legacy of the systemic violence of its post-colonial past, the migrant labour system for mines, and the recent history of racial segregation. Rapid urbanisation has also led to the development of large urban slums that lack the requisite physical and social infrastructure to build social cohesion, with easy access to cheap alcohol.(17) Socially, violence has been normalised as a frequent feature of civil protest and political discourse, and the hegemonic form of masculinity is patriarchal. South Africa also has high levels of legal and illegal firearm ownership and the highest rates of incarceration – an additional exposure to institutional and interpersonal violence – in Africa, where the ratio of male to female prisoners is double the global average.(18) It is not surprising that this is a society in which interpersonal violence is expected, and that the forms that it takes are highly gendered. Common reactions to adverse events tend to differ between men and women. Men are socialised into coping by externalising through anger, irritability, violence against intimate partners and others, and increased engagement in risk-taking behaviours.(19) This, alongside the high levels of violence to which males are exposed across the life course,(20) engenders a continuous, and often intergenerational cycle of violence.

South African data have consistently shown that men are not only the main perpetrators of violence,(5) but that men also have an overwhelmingly higher risk of violent death. Part of this may relate to prevailing gender norms in which men identify with the role of “protector”.(21) Defending honour and asserting dominance over others may increase men’s resistance in the face of conflict, in turn increasing the risk of fatal outcomes. This was shown in a recent South African study of co-occurring violence during robbery events in which male victims were significantly more at risk of a fatal outcome.(22)

Violence against women is endemic in South Africa, with rates almost six times global rates. South Africa has responded proactively to such violence with interventions and policy measures culminating in a National Strategic Plan on Gender-based Violence & Femicide, including measures to strengthen the criminal justice system, promote accountability across the state and support survivors. However, men’s disproportionate burden of homicide has not resulted in targeted, meaningful prevention.

Whereas the number of female homicides decreased over time, the number of male homicides, and hence their share of all homicide, increased from 2009 to 2017.(2) Yet this has not changed the prevailing socially normative perception that men are neither vulnerable to, nor the victims of, trauma.(20) Ratele et al (2016) suggest that this limited engagement with evidence of men’s vulnerability has inadvertently pathologised black males in South Africa, and prevents us from recognising that boys and men are legitimate recipients of violence prevention interventions.(23) There is an urgent need for effective interventions that target men and address not only the gender norms that increase risk, but also the structural drivers of homicide that are rooted in poverty and socio-economic inequality.

In comparison with global averages, South African men, women and children were all subject to abnormally high levels of homicide risk. Age-standardised homicide rates were similar in 2017 and 2009.(2) However, the share of overall mortality due to homicide – ranked 8th and accounting for 3.5% of all-cause mortality in 2012(3) – is set to rise as mortality from other major causes such as HIV continues to decrease,(24) while the share of homicide among all injury deaths is expected to rise due to a decrease in road deaths.(7) The global decline in homicide has also resulted in the relative risk of homicide in South Africa increasing from 5.8 to 7.1 times the global average from 2009 to 2017.

The male to female ratio was highest for firearm discharge, the second leading cause. The implementation of the Firearms Control Act was associated with reductions in firearm homicide, but poor enforcement is associated with a subsequent surge in gun deaths.(25) Males, who are also more likely to be armed,(22) accounted for the larger share of the increase in gun deaths reported between 2009 and 2017. The higher incidence of homicide on weekends (and holidays) affirmed the prominent role of alcohol, present in 55% of male and 38% of female homicide cases. South Africa’s drinking pattern is characterised by very high levels of heavy episodic drinking, particularly amongst males, which is reflected in the gender distribution of homicides attributable to alcohol. Males accounted for 95% of the estimated 15,168 alcohol attributable-homicides in South Africa in 2000, 2006 and 2012.(26) Alcohol sales bans implemented alongside lockdowns during South Africa’s COVID-19 pandemic were associated with reductions in non-natural deaths and trauma cases,(27,28) but this critical policy window has not, as yet, translated into more sustained approach to reduce alcohol harms.(29)

There was considerable interprovincial variation in overall homicide. While the homicide risk was higher for men than women in all provinces, the provinces with the highest male homicide rates also ranked highest for female homicide. This suggests violence is endemic in some provinces, which will require complex population level approaches to prevention that address social determinants and norms that support violence. The temporal pattern, with the highest homicide incidence in months that coincided with school holidays and the year-end and easter festive seasons, rather than the warmer months, did not support the theory that aggression (and with it homicide) is associated with increases in temperature.(30) The temperature range from winter to summer in South Africa may be insufficient to affect aggression levels, but further analysis should explore the potential confounding effect of alcohol consumption on the temporal pattern.

South Africa is one of few countries worldwide with a quadruple disease burden where injuries feature alongside major infectious and non-communicable diseases and the largest HIV pandemic worldwide. The burden is compounded by the deleterious effects of violence on other outcomes - mental health and developmental issues such as substance abuse, chronic conditions (e.g. gastrointestinal, gynaecological and fatigue-related), absenteeism and loss of work - and, moreover, is a major impediment to social development.(31,32) Yet despite its importance among major causes of injury, there is no sense of urgency about addressing violence as a structural issue, and interventions and policies to reduce population-level homicide have been ineffective.

With male violence frequently located in the public space,(22) there is potential to converge the prevention agenda to reduce male and female homicide jointly, which might give population-level approaches more impetus. This would also align with the Sustainable Development Goals (SDGs), which have substantially expanded the scope of violence prevention by advocating the reduction of ‘all forms of violence everywhere’ (Target 16.1), eventually – but only implicitly - recognising men as potential targets for prevention action. The urgent need to address violence against women and children should therefore be integrated into an inclusive approach to address violence in line with SDG16.(33)

There remains an urgent need for homicide and other violence indicators to be disaggregated by sex, to shed further light on socio-demographic-specific risk factors and situational pathways to homicide in violence of various types. This is not only the case in South Africa. The WHO explains the critical importance of disaggregating data for health systems, in particular to provide information to allocate appropriate resources; yet the WHO itself only started reporting disaggregated global health statistics in 2019. The forthcoming male homicide study, the second phase of the current study, will be the first research to profile male victims and perpetrators.

The study has several limitations. The sample size was adequate for estimating population incidence rates in 2009 and 2017 at the national level, but the study lacked power to compare rates for certain subgroups between study years. In addition the sampling frame was different between the two years, with a smaller sampling frame in 2009, devised to compare injury rates across metropolitan and non-metropolitan populations, but limiting our ability to compare provincial homicide patterns. Another limitation was that with only two time points we could not test for trends in male homicide rates.

Despite these limitations, our study demonstrates the value of mortuary-based survey data for estimating sex-disaggregated homicide data in the absence of routine injury mortality surveillance and confirms that this approach is feasible in a high violence resource-limited setting.

## PUBLIC HEALTH IMPLICATIONS

Our study highlights the extraordinarily high levels of homicide in South Africa, the disproportionate burden borne by adult men, and the negligible response to date. We urgently need a redoubling of efforts to control alcohol and firearms, which have already been shown to influence rates of violence in South Africa, as well as programmes to address the insidious effect of societal norms that drive the excessive burden of physical violence borne by men, and structural interventions to overcome the root causes of poverty and inequality. We consider our study to be an important and necessary – if belated - first step to identify specific groups at increased homicide risk who could benefit from specific interventions and policies. Only through challenging the notion of male invulnerability can we begin to address the enormous burden of violence born by men.

## Data Availability

All data produced in the present study are available upon reasonable request to the authors.

## Acknowledgements

We are grateful for the support of forensic pathology services across all provinces and, in particular, to Professor Emeritus Gert Saayman of the Faculty of Health Sciences, Dept of Forensic Medicine, University of Pretoria; Prof Jeanine Vellema, the Retired Head of the Clinical Department of the Gauteng Department of Health Forensic Pathology Service Southern Cluster and an Honorary Member of the University of the Witwatersrand Division of Forensic Medicine & Pathology; and Dr Sibusiso Ntsele of the eThekwini Forensic Pathology Services, KwaZulu-Natal Department of Health, Durban, South Africa.

