## Supplemental Table 1 for "South Africa’s male homicide epidemic hiding in plain sight: exploring sex differences and patterns in homicide risk in a retrospective descriptive study of postmortem investigations": Supplementary Table 1.docx (from another version of this preprint: version 1, 2023-02-04)

**Supplementary Table 1. External cause of homicide categories included in the injury-related mortality survey and corresponding ICD-10 codes, South Africa, 2017**

| **Cause of homicide** | **ICD-10 code** |
| --- | --- |
| Poisoning | X85 - X90 |
| Strangled/asphyxiated/suffocated | X91 |
| Drowning/ immersion | X92 |
| Firearm discharge, no | X93 - X95 |
| Fire /other burn | X97, X98 |
| Sharp force | X99 |
| Blunt force | Y00, Y04 |
| Pushed from a height | Y01 |
| Neglect and abandonment* | Y06 |
| Assault by other specified means | Y08 |
| Assault by other unspecified means | Y09 |
| Legal intervention** | Y35 (Y35.0-Y35.7) |

* Applied exclusively to new-born children.

** Applied as a secondary code in addition to the primary codes listed above.
